# Current state of COVID-19 knowledge, attitude, practices, and associated factors among Bangladeshi food handlers from various food industries

**DOI:** 10.1101/2021.01.06.21249118

**Authors:** Md. Fahad Jubayer, Md. Shahidullah Kayshar, Md. Faizul Kabir, Md. Shoaib Arifin, Md. Tariqul Islam Limon, Md. Nasir Uddin, Md. Al-Emran

## Abstract

While people around the world are terrified of the global pandemic coronavirus disease 2019 (COVID-19) and are dying for a permanent solution, undertaking preventive safety measures are said to be the most effective way to stay away from it. People’s adherences to these measures are broadly dependent on their knowledge, attitude, and practices (KAP). People working in the food industries must be extra cautious during this time because they are in close proximity to consumable items. The present study was designed to evaluate food handlers’ knowledge, attitude, and practices regarding COVID-19 in different food industries in Bangladesh. A number of 400 food handlers from 15 food industries took part in this online-based study. The information was collected from the participants through a questionnaire prepared in Google form. With a correct response rate of about 90% on average (knowledge 89.7%, attitude 93%, practices 88.2%), the participants showed an acceptable of KAP regarding COVID-19. Education and working experiences had a significant association with the total KAP scores (p < 0.05). The findings may assist public health professionals and practitioners in developing targeted strategies for implementing such studies in other industrial sectors and taking appropriate measures based on the KAP studies.

## 1. Introduction

Coronavirus disease 2019 (COVID-19) is considered as one of the most widespread pandemics since long. It has become a worldwide public health threat due to its high transmission rate and unusual infection characteristics. After the first case being diagnosed in Wuhan, China, in December 2019, the world health organization (WHO) declared it a global health emergency. As of September 12, 2021, there are more than 220 million confirmed cases in almost every country and territory with a burden of nearly 5 million deaths (WHO, 2020). This disease has already made its deadly mark in Bangladesh since the first case spotted on March 08, 2020. In Bangladesh, the most confirmed cases of COVID-19 are in Dhaka, with almost 30% of total cases across the country (IEDCR, 2020).

Bangladesh is still in a grave condition where thousands of people are getting affected every day. All educational institutions are still remained closed, although online classes and lectures are going on (Jubayer et al., 2021). The government offices are also functioning, maintaining all safety measures. The economy of this country is mostly reliant on the processing and manufacturing sectors, and most people make their living through daily income. Nevertheless, ‘working from home’ is not a feasible option for food processing or any other manufacturing industry (Maria et al., 2020). Also, for avoiding the risk of food shortage, the highest priorities should be given on supply chains to function appropriately (Galanakis, 2020). That is why it is necessary to ensure good health and safety for the food handlers providing high importance in maintaining physical distance in manufacturing plants (Shahidi, 2020). According to the U.S. Department of Homeland Security, a food processing industry is a critical infrastructure, where the workers are needed to work in an enhanced safe environment (Dyal, 2020)

Along with food safety, the manufacturers should now encompass measures for the prevention and control of coronavirus within their plant (SGS, 2020). The SARS-CoV-2 is not a foodborne disease, as there is no evidence of transmission through any food product (EFSA,2020; Shahidi, 2020). Standard handling procedures for food is supposed to be enough for the management of COVID-19 inside the factory, so long as personal distancing and additional safety procedures are conformed (SGS, 2020). However, no respiratory virus is transmitted via food. But, there is a chance of transmission if any infected person touches the food, sneezes or coughs on it, and after that, another person makes contact with the same food and touches his nose, eyes, or swallows it (Galanakis, 2020; Rizou et al., 2020).

Therefore, careful handling of foods and packaging materials, practicing proper personal hygiene, especially washing and sanitizing hands extensively, should be ensured by the management of every food manufacturing industry. The FDA has suggested cleaning and sanitizing the food contact surface before production operation as precaution measures (Seymour et al., 2020). So, it may be possible for the workers to be infected with the virus through contact surfaces and surroundings when working.

For successful control measures of this coronavirus disease (COVID-19), the people must obey the control measures as suggested by the World Health Organization (WHO, 2020). The success of implementing preventive measures is mostly dependent on the knowledge, attitude, and practices of an individual (Ajilore et al., 2017; Karim et al., 2020; Zhong et al., 2020). Several studies revealed that the prevention of the previous SARS outbreak was associated somewhat with the knowledge and attitude of people towards infectious diseases (Tao, 2003; Person et al., 2004). However, after nearly two years of the pandemic’s onset, there are very few authentic studies or reports (Jubayer et al., 2020) on food handler safety, food safety, and measures taken in Bangladesh’s food industries during this time. Also, consumer health, worker’s health, and working environment are of paramount concern. To enable and facilitate COVID-19 management within Bangladesh’s food industries, we feel compelled to comprehend and assess whether the current state of workers’ awareness and knowledge of the disease at this critical juncture is sufficient or if additional steps are required to bring it to a satisfactory condition. As a result, the current study was conducted to investigate the current state of COVID-19-related knowledge, attitude, and practices (KAP) and the associated factors influencing the KAP status of food handlers from various food industries throughout Bangladesh.

## 2. Methods

### 2.1 Study Place and Population

A cross-sectional study was carried out among 400 food handlers in fifteen (15) food industries all across Bangladesh between November 20, 2020 and February 20, 2021. Participants in this study were the general floor workers. We selected people from the production, quality assurance (QA)/ quality control (QC), store, maintenance, and human resource management (HRM) departments to participate in this survey. In most of the industries, the average production time was 8 hours per day rather than 24 hours, while all possible safety measures were maintained. The inclusion criteria for the study were, a) being a Bangladeshi resident, b) having internet access, and c) participating voluntarily.

### 2.2 Procedure

For avoiding close contact with the respondents, we decided to design a web-based questionnaire. A structured questionnaire (Bengali and English) was prepared online (Google forms), and the link was sent to the respondents through email and social media (WhatsApp and Facebook messenger). A concise and straightforward completion guideline was displayed at the beginning of the questionnaire. In the first part, respondents were required to fill information designed to gather demographic data. The second part was on the knowledge, attitude, and practices regarding COVID-19. The knowledge part was divided into two parts, namely, general COVID-19 knowledge (10 questions) and knowledge related to food safety (10 questions). The attitude and practices part was comprised of 10 questions each. Questions from the KAP section were answered as yes/no, agree/disagree, yes/no basis, respectively. A correct and desired response received one point, and the wrong or undesired answer received zero point. For the determination of adequate knowledge (>80%, score>16), positive attitude (>80%, score>8), as well as desirable practices (>80%, score>8), Bloom’s cut-off point of 80% was utilized (Kaliyaperumal, 2004). Cronbach’s alpha was 0.75 for the questionnaire, which indicates that the questionnaire is a valid and reliable instrument for the purpose of assessment.

### 2.3 Statistical Analysis

Various tabular and statistical analyses have been used to conceptualize the whole scenario. SPSS version 16.0 was used for statistical analysis. The primary analysis of data showed non-normality in the sktest and Shapiro-Wilk test; hence we have used nonparametric alternative tests, namely, Mann-Whitney U test, Wilcoxon W test, and Kruskal-Wallis H test. KAP scores of respondents are presented as mean and standard deviation. Categorical variables are computed as percentages. A linear regression model has been performed to determine underlying factors that enhanced KAP scores. In the regression model, gender is a dummy variable; age and experience are continuous variables, and the education and work area belong to categorical variables.

### 2.4 Ethical Consideration

The ethical committee of Sylhet Agricultural University approved the study. Respondents were also asked to provide informed consent. During the study, however, no personal information was used. All data were kept on a password-protected computer that was only accessible to the research team.

## 3. Results

### 3.1 Demographic Characteristics

The demographic characteristics of participants are shown in **Figure 1**. The mean age of the respondents was 32.9±8.5, with the majority being male. Most of them were aged between 30 and 40. Almost everyone (93.5%) had some level of education, among which the maximum percentage (41.7%) was in the secondary education group. The rest had university and technical education. The majority of respondents (46.2%) had working experience between 2 and 5 years, while the least (10.5%) had working experience of more than ten years. Most of the food handlers among the participants were from production department followed by QA/QC, maintenance, store, and HRM. While no one was found unaware of the situation, it appeared that a significant number of people (54.7%) acknowledged social media as a source of information regarding COVID-19.

**Figure 1.**
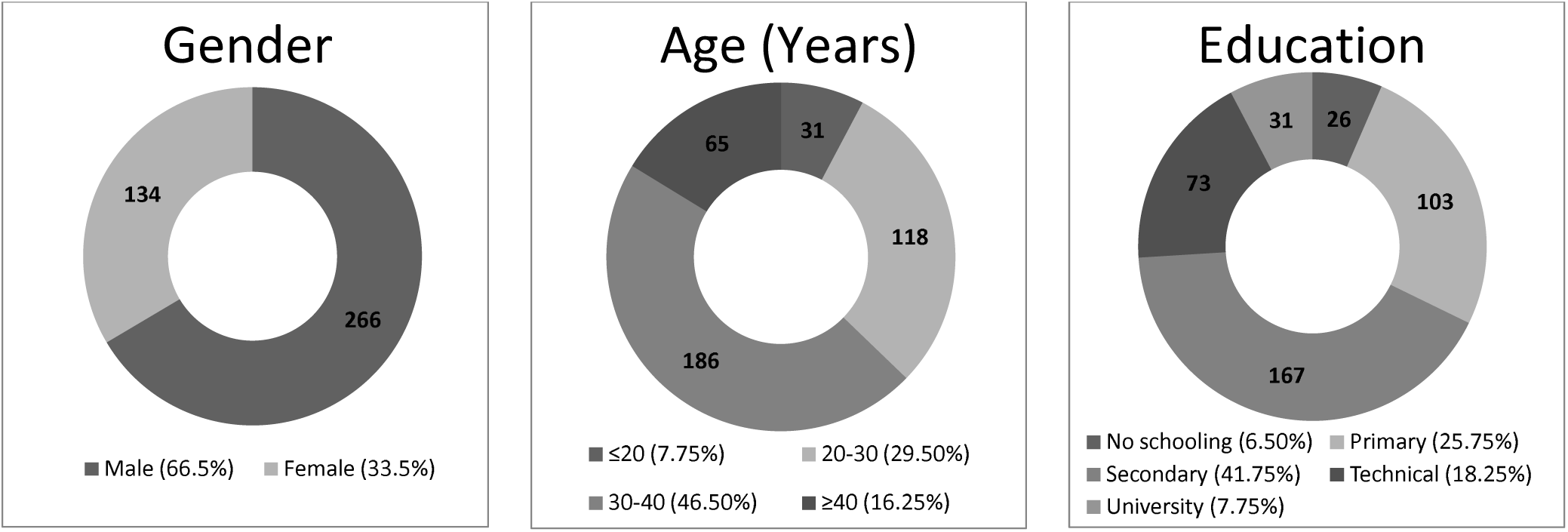
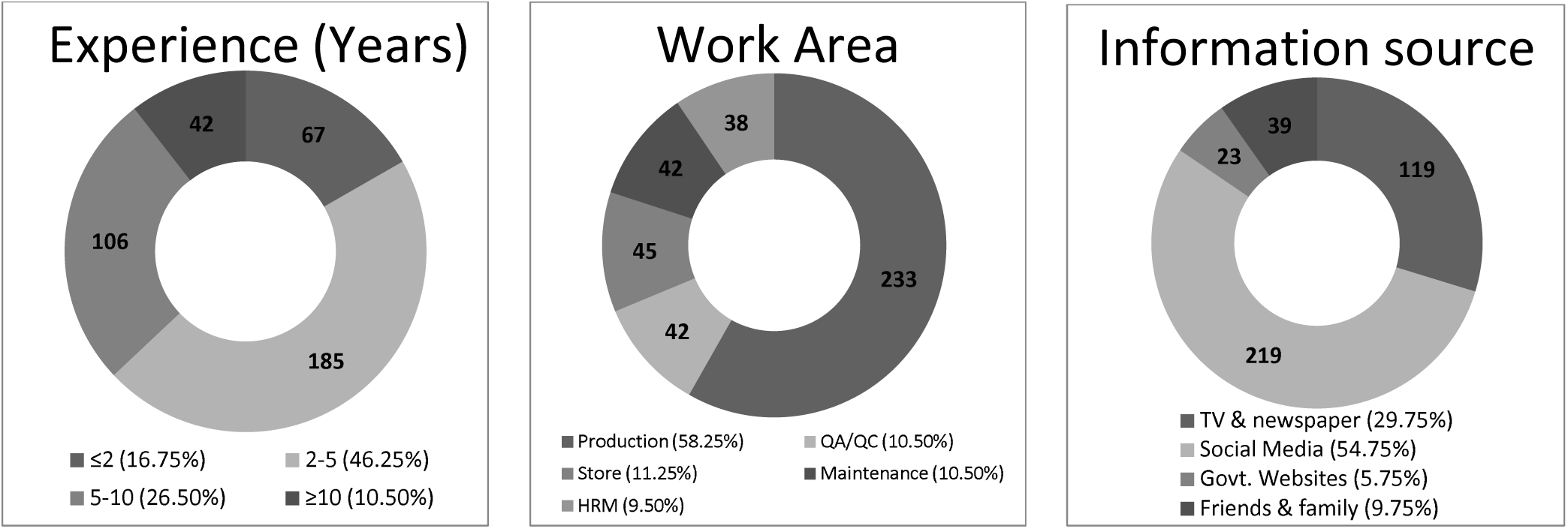
Demographic characteristics of respondents.

### 3.2 Knowledge, Attitude, and Practices of Respondents

About 89.7% of respondents provided the correct answer with a knowledge score of 17.9±0.08 (**Table 1**). Questions about general COVID-19 knowledge received 91.6% accurate responses, while questions concerning food safety received 87.9% correct answers. They had sufficient knowledge on most of the issues. But when it came to the survival and destruction condition of the coronavirus, they seemed to have some problems. Whether the virus can survive at freezing temperatures or whether UV can kill it got less than 80% correct responses. About two out of every ten workers were unable to answer questions about comorbidity risks or zoonosis and were unaware of any registered medication on the market.

**Table 1.** Knowledge of respondents regarding COVID-19.

| Knowledge |  | Correct responses<br>n (%) |
| --- | --- | --- |
| General | 1. SARS-CoV-2 is a disease caused by virus. | 384 (96.0) |
|  | 2. Fever, cough, sore throat, breathing problems are common symptoms of this disease. | 377 (94.3) |
|  | 3. COVID-19 can be fatal. | 387 (96.8) |
|  | 4. COVID-19 can be caused by close contact with infected person. | 395 (98.8) |
|  | 5. COVID-19 can spread via respiratory droplets. | 382 (95.5) |
|  | 6. Patient with diabetes, CVD, and asthma are at a higher risk of infection. | 348 (87.0) |
|  | 7. Are there any registered medicines available in the market? | 335 (83.8) |
|  | 8. This disease cannot be transmitted to human from pet animals. | 331 (82.8) |
|  | 9. This disease can be spread in hot and humid climate. | 374 (93.5) |
|  | 10. Antibiotics are effective against COVID-19. | 351 (87.8) |
| Food safety | 11. SARS-CoV-2 can grow on baked product. | 395 (98.8) |
|  | 12. The virus can stay on the packaging material for long time. | 361 (90.3) |
|  | 13. This virus can survive at freezing temperature. | 297 (74.3) |
|  | 14. Viruses cannot multiple and increase in numbers in foods. | 333 (83.3) |
|  | 15. Cooking is the only way to be sure that foods made with flour and raw eggs are safe. | 371 (92.8) |
|  | 16. Heating foods (70°C for 2 minutes or equivalent) is required to kill pathogens. | 359 (89.8) |
|  | 17. Alcohol-based disinfectants can effectively reduce infectivity of SARS-CoV-2. | 389 (97.3) |
|  | 18. UV is effective against COVID-19. | 309 (70.3) |
|  | 19. Pasteurization can kill SARS-CoV-2. | 281 (85.3) |
|  | 20. SARS-CoV-2 can spread by water. | 387 (96.8) |

With a score of 9.3±0.04, more than 90% of people expressed a positive attitude towards different COVID-19 issues (**Table 2**). More than a few of 10% of participants did not think they could be affected while working and nearly the same number did not want to be trained on COVID-19. Nearly 13% didn’t believe that washing one’s hands for at least 20 seconds was required.

**Table 2.** Attitude of respondents towards COVID-19.

| Attitude | Positive attitude |
| --- | --- |
|  | n (%) |
| 1. I can get this disease while working in the factory. | 359 (89.8) |
| 2. Washing hand for at least 20 seconds is necessary to prevent transmission. | 351 (87.8) |
| 3. It is necessary to report your illness or any symptoms to the management immediately. | 371 (92.8) |
| 4. I think I need proper training on COVID-19 for ensuring personal safety. | 356 (89.0) |
| 5. I am worried of my family members as I am working outside. | 392 (98.0) |
| 6. Is it necessary to avoid coming to work if you feel any sickness. | 374 (93.5) |
| 7. A COVID-19 patients should be kept in isolation for certain period. | 385 (96.3) |
| 8. Use of mask can prevent contamination. | 387 (96.8) |
| 9. Maintain safety measures outside the factory are also important. | 384 (96.0) |
| 10. Factory should remain closed until the condition gets better. | 361 (90.3) |

Food handlers demonstrated desirable outcomes in their practices also (**Table 3**). On average, about 88% of workers maintained commendable and appropriate practices during the pandemic. However, some issues, like staying away from crowded places, not touching eyes, nose, or mouth during food handling, and avoiding frequent use of public transportation, were not regularly practiced by nearly 20% of the workers.

**Table 3.** Practices of respondents during COVID-19.

| Practices | Desirable practices |
| --- | --- |
|  | n (%) |
| 1. Do wash and sanitize your hands more frequently than before? | 369 (92.3) |
| 2. Do you update yourself regularly about the situation? | 338 (84.5) |
| 3. Do you touch your eyes, nose or mouth while handling food and food packages? | 315 (78.8) |
| 4. Have you gone to any crowded place in last two week? | 299 (74.8) |
| 5. Do you wear mask outside the factory? | 350 (87.5) |
| 6. Do you usually shake hands with others after the breakout of COVID-19? | 385 (96.3) |
| 7. Do you touch foods in empty hands in the production line? | 394 (98.5) |
| 8. Do you change your mask after coughing and sneezing inside the factory? | 377 (94.3) |
| 9. I discuss the prevention measures and make aware my family members of COVID-19. | 364 (91.0) |
| 10. I try to avoid public transportation. | 335 (83.8) |

Scores on knowledge, attitude, and practices have been tested by different demographic characteristics, i.e., gender, age group, education, experience, work area in current company, and source of information on COVID-19 (**Table 4**). The null hypothesis of the Mann-Whitney U and the Wilcoxon test stipulates that each group’s distribution was the same. In the case of knowledge, all demographic characteristics showed significant differences in central tendencies for each distribution. When it comes to attitude, all demographic characteristics other than the job area showed significant differences. Education was the only factor that showed a significant difference in the test for practices score. Therefore, the demographic characteristics lead to varying the individual score, while education possesses significant results in all three KAP elements.

**Table 4.**
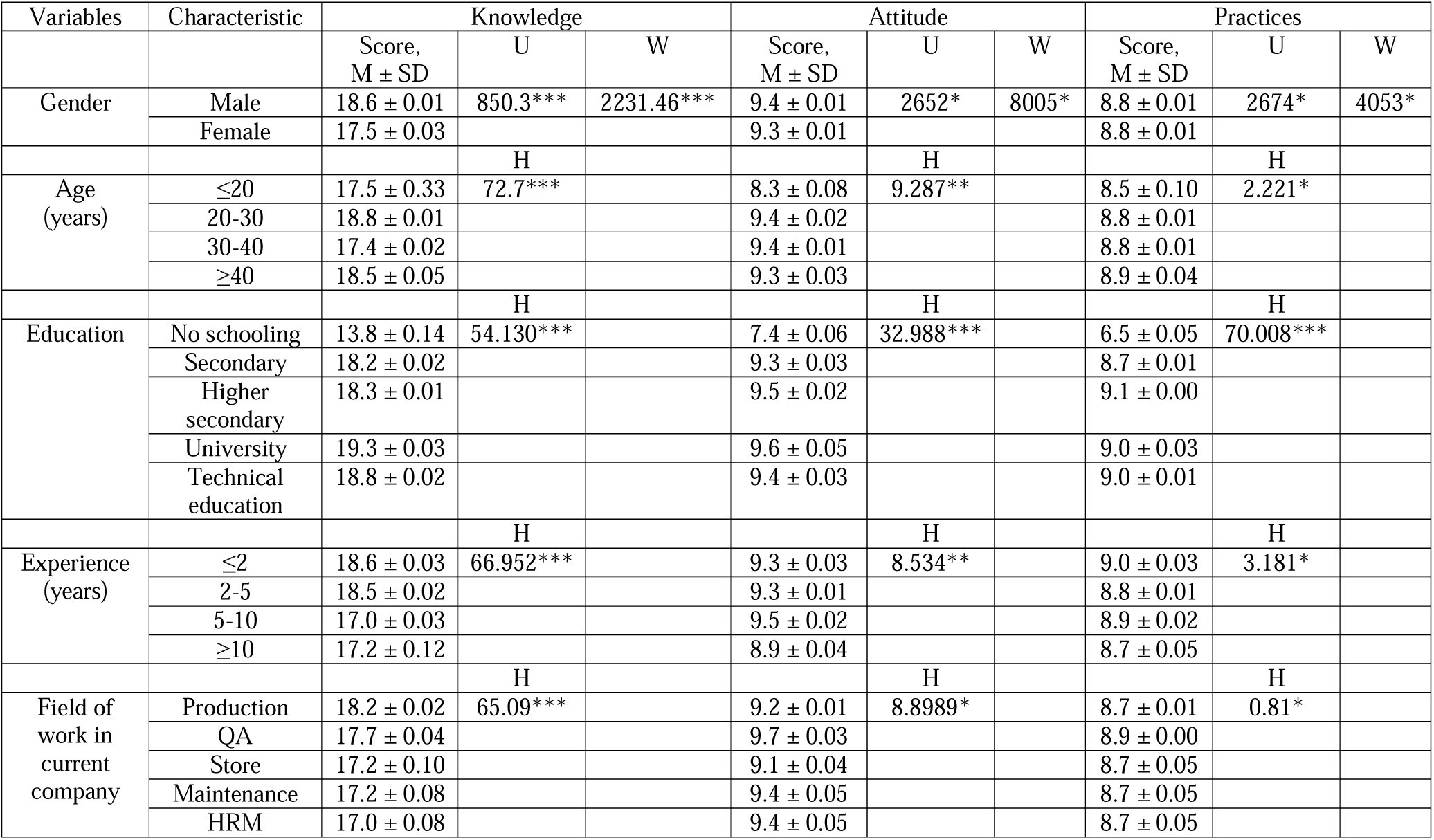
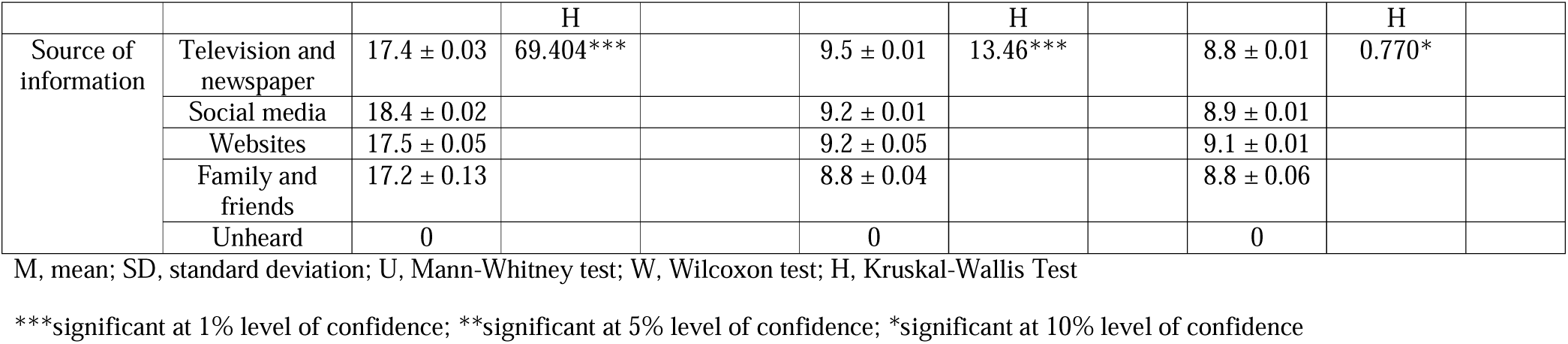
Differences in KAP scores according to the demographic characteristics.

Other than the individual score on KAP elements, in the regression model, we have determined underlying factors that enhanced the total KAP score. The R-square value indicates the global check for the model that the model explains 81.4% of the variance in total score on knowledge, attitude, and practices (**Table 5**). F value is very close to zero, which indicates that the regression model is statistically significant and predicts the outcome variable. In this regression model, education and experience significantly associate the knowledge and practices of COVID-19. Age, gender, and work area have no significant association with the knowledge, attitude, and practices towards COVID-19. Those who attended primary school have 7.5 times more knowledge of COVID-19 than those who did not attend school, and it is 6.8 times higher for those attended a high school, 7.5 times higher for those with a university degree, and 6.6 times higher among technical education holders than non-school-attending participants. Experience is also statistically significant at a 5% level of confidence, but unexpectedly the coefficient is negative in value, although very minimal. That shows that those in the profession who are younger are more cautious or knowledgeable about COVID-19 than the older employees.

**Table 5.** Regression of total KAP scores.

|  |  | Coefficient | p-value |
| --- | --- | --- | --- |
| Constant |  | 21.2*** | 0.00 |
| Gender |  | -0.2*** | 0.6 |
| Age |  | 0.1** | 0.4 |
| Education | (Base-No Schooling) |  |  |
|  | Primary | 7.5*** | 0.00 |
|  | Higher Secondary | 6.8*** | 0.00 |
|  | University | 7.5*** | 0.00 |
|  | Technical Education | 6.6*** | 0.00 |
| Experience |  | -0.6** | 0.01 |
| Field of work | (Base- Production) |  |  |
|  | QA | -0.1 | 0.7 |
|  | Store | -0.03 | 0.8 |
|  | Maintenance | -0.004 | 0.8 |
|  | HRM | -0.6*** | 0.3 |
| R-Square | 81.4% | F-value | 155 |
| Prob>F | 0.00 |  |  |

\*\*\*significant at 1% level of confidence; \*\*significant at 5% level of confidence
p-value < 0.05 means we can reject the nulls, hence for both cases the central tendencies of both distributions differ.

## 4. Discussion

Since the spring of 2020, there have been numerous Covid-19 outbreaks in Europe and around the world among food industry workers. Tönnies, one of Germany’s largest meat processing companies, for example, sparked one of the most heated debates on the subject after it was revealed that many of its workers had contracted the coronavirus. Over 1,500 people have tested positive for COVID-19 so far. Many meat processing companies in EU member states like Belgium, France, Ireland, Spain, Poland, and the Netherlands have seen coronavirus outbreaks at their plants since April 2020. A total of 950 laborers contracted COVID-19 at 19 plants in a meat factory in Ireland. At some plants, one-fourth of the entire workforce fell ill. Two French slaughterhouses reported virus outbreaks in mid-May. So far, 180 laborers have tested positive for the virus (EFFAT, 2020).

From the perspective of Bangladesh, a densely populated and lower-middle-income country, social distancing is visibly impossible. People instead work outside in this dangerous situation than starving (Anwar et al., 2020). Food and beverage industries worldwide are likely to experience variable impacts of COVID-19 on different stages of its value and supply chain (Manepalli & Nagvenkar, 2020). The food industries of Bangladesh are no exception. Right now, primary concern for the food industry is to provide all support to keep their employees healthy and available (Foodware365, 2020). Food handlers are usually exposed to various biological materials (Peng et al., 2020). Moreover, the coronavirus can persist in the food industry’s regular handling surface, e.g., metal, plastic, and glass, for a couple of days (Kampf et al., 2020). Therefore the workers need to have knowledge of proper safety rules and bear attitude for practicing those to stay safe from spreading COVID-19.

To the best of our knowledge, this current study is the first that evaluates the KAPs of food handlers of food industries towards the pandemic COVID-19. However, since the outbreak, we found some studies regarding KAPs and COVID-19 conducted with healthcare workers and physicians (Ahmed et al., 2020; Huynh et al., 2020; Khader et al., 2020; Olum et al., 2020; Saqlain et al., 2020; Shi et al., 2020; Zhang et al., 2020), students (Taghrir et al., 2020), and general people (Abdelhafiz et al., 2020; Azlan et al., 2020; Karim et al., 2020; Roy et al., 2020; Zhong et al., 2020).

In the present study, the respondents from these food industries had sufficient knowledge regarding COVID-19. Almost 90% of them provided a correct response in different questions asked on COVID-19. The knowledge section was divided into two parts, namely, general and food safety, that was responded correctly by 91.6% and 87.9% of participants, respectively. Similar kind of performances where more than 80% respondents have accurate knowledge on COVID-19 can be found in some recent studies by (Saqlain et al., 2020) (93.2%), (Zhong et al., 2020) (90%), (Shi et al., 2020) (89.5%), (Zhang et al., 2020) (89%), (Taghrir et al., 2020) (86.9%), (Olum et al., 2020) (82.4%). All these industries have different certifications, such as ISO, HACCP, HALAL, and so on, and they need to arrange food safety training for all of their employees on a regular basis. This may be one of the reasons for their performances in food safety knowledge. Although it is not possible to compare the results to any other studies on the food industry and COVID-19, we discovered that the food safety knowledge level of food handlers from various food industries was satisfactory (Ansari-Lari et al., 2010; Adesokan & Raji, 2014; Jianu & Goleţ, 2014).

The respondents seemed to have well acknowledged about the necessary information on COVID-19. The causative organism, symptoms, severity, and consequences of the infection, how it spreads – are usually known to almost 9 in every ten people. Comparatively, few people were aware of whether this disease can transmit from pet animal to human and if there is any registered treatment option or medicine available in the market. Due to the spreading of panic and fake news in this pandemic time, many owners and people fear that pets might transmit the disease to them. There is no significant proof of substantial threat from pets or animals (Parry, 2020). More than 10% of them were not sure that diseases like diabetes, CVD, and asthma could be deadly in this situation. Usually, participants with no education appeared to have very poor knowledge of this issue. People who acquire information about COVID-19 from their friends and family were also a bit unaware of this matter. Some workers did not have education, solvency, and had no access to the internet and smartphones. They mostly depend on other people for any information on COVID-19. As a result, they were unaware of several critical pieces of information.

When we asked them questions regarding the COVID-19 and food safety, correct response percentages (87.9%) were lower than the general part (91.6%). More than 25% failed to answer the question regarding the survival of SARS-CoV-2 in freezing temperatures. According to the WHO, this organism can survive at −20 °C for nearly two years, and transmission via frozen food can be possible (WHO, 2020). A study found that among people from different occupations in Bangladesh, there is a significant knowledge gap about the incubation and transmission characteristics of this virus (Mannan & Mannan, 2020). When it comes to relating the freezing temperature and microorganisms, several studies demonstrated a low level of food handler’s knowledge regarding this (Bas et al., 2006; Galgamuwa et al., 2016; Akabanda et al., 2017; Al-Kandari et al., 2019; Ansari-Lari et al., 2020). But the level of performance and knowledge level on related topic of the respondents in current study is undoubtedly better than these studies.

Usually, for the inactivation of viruses, a typical cooking temperature of more than 70°C is sufficient (für Risikobewertung, 2020). The workers knew that cooking and heating is enough for the product to make it safe from pathogens. Besides, almost everyone (97.4%) agreed that alcohol-based disinfectants could be effective against SARS-CoV-2. The SARS-CoV-2 cannot spread through drinking water. Commonly used treatment methods are competent enough to make the water safe (CDC, 2020), and it was known to almost all workers. Besides, cleaning of processing surfaces and conveyors with disinfectants is an everyday job for the floor workers. Therefore, dealing with the questions on thermal processing and basic hygiene became easy for them. Whether the virus can survive at freezing temperature or, can the ultraviolet rays destroy it – is unknown to many respondents. But, a fair amount of workers answered correctly in question regarding the effect of pasteurization on the coronavirus.

From the present study, we found that majority of respondents had a desirably positive attitude towards the COVID-19 issue. Except three of them, all said that they remained worried about their family members during this outbreak. Most people working in different processing and manufacturing industries are from outside the city and leave their families in their hometown or village (BBS, 2017). Furthermore, due to transportation lockdown, it was hard for them to visit their family. Why the three participants declared of not being worried about family was unknown to us. About 10% thought that the industries should run its operation in the time of COVID-19, and they will not get infected while working in the factory. Besides, about 7% of them wanted to come to work although they fell ill, and 8% of the workers did not want to let the management know about their sickness. Although not many in numbers, these few peoples can cause a threat to other healthy people during the time of work if they are infected with the coronavirus. In a country like Bangladesh, workers’ socioeconomic conditions and challenges may drive them to work while feeling sick. During the lockdown, a significant number of people have lost their jobs in Bangladesh due to the closure of industries. Many industries cut salaries while some were unable to pay the incentives before the Eid festivals, 2020 (Amnesty International, 2020). More than 10% of participants did not agree on washing hands for at least 20 seconds. Perhaps, they thought maintaining a particular time was not necessary. Respondents from the no education group and those who said to collect information from family and friends were mostly disagreed on this issue.

Most of the respondents (95% on average) reported washing and sanitizing hands more often, avoiding shaking hands and touching foods while working in a line, and altering masks after coughing and sneezing. This indicates the frequent changes and awareness of workers for preventing the COVID-19. Surprisingly, 7 in 10 participants said they usually made contact with eyes, mouth, and nose while handling food products in the production line. It is essential to know the appropriate use of gloves and masks in the food industry while handling the products (WHO, 2020). This kind of unexpected behavior of not washing or sanitizing hands and touching face, nose, and eyes after coughing or sneezing can also be seen in a study (Prabhusaran et al., 2018). We want to say that the participants know the risks of touching the face during this time, but such strict practices may take some more time. Also, there is a chance of considering themselves less likely to the transmission, which may be stated as ‘optimistic bias’ (da Cunha et al., 2014).

A remarkable number of people (25.2%) found to go outside more than once in the last seven days, and about 13% of total respondents were not used to wear masks outside the factories. Moreover, some of them use public transport regularly. This kind of practice could lead a person to transmit more often, which would make all other employees prone to infection within a week or two. This kind of occurrence can force the business to close immediately (SGS, 2020). In another study from Bangladesh by Haque et al., (2020), it was observed that nearly 30% of the participants frequently go outside and meet their friends. On the contrary, many improved practices in avoiding crowds can be seen in different study groups from Malaysia (Azlan et al., 2020), China (Zhong et al., 2020), and Iran (Taghrir et al., 2020).

According to the study, the total KAP scores were not affected by gender, age, and working sections, although there were differences in the knowledge scores. We found that the males, workers of 20 to 30 years old, and those working in the production departments significantly have higher knowledge scores. When, in most of the recent KAP studies (Azlan et al., 2020; Abdelhafiz et al., 2020; Neamti et al., 2020; Olum et al., 2020; Zhong et al., 2020) female participants showed higher scores than male participants. In different studies, people of different age limits showed different outcomes in knowledge scores. This is hard to compare these results with the food industry people as they have separate working environments. For example, respondents above 40 and 50 years old, respectively, had superior knowledge compared to other age groups (Azlan et al., 2020; Huynh et al., 2020). Another study found that respondents between the ages of 18 and 30 performed better on the COVID-19 knowledge-related questions (Abdelhafiz et al., 2020).

The exciting thing is that the respondents with less experience are superior in their total KAP scores. The coefficient is negative in value in case of total working experience (**Table 5**). Therefore, the younger food handlers have good knowledge of COVID-19, and they are positive in their attitude and approach to this pandemic than the elderly participants. It is a novel virus, and the young have more access to the internet and other information due to being technologically sound. A similar kind of result was observed by (Huynh et al., 2020). Contradictory results can also be seen in a study, where participants with more experiences had a higher level of knowledge (Nemati et al., 2020). Education showed to have a significant role in the KAP scores of the participants. Those who have completed graduation are superior in KAP. This finding is in line with the studies of (Abdelhafiz et al., 2020; Olum et al., 2020; Zhong et al., 2020).

In the current study, most food handlers reported different social media as their sources of information on COVID-19. A fair number of people referred to television and newspapers for the latest update, followed by those who depend on websites, family members, and friends. Those who kept themselves updated through social media had significantly better knowledge than others. However, social media followers lagged behind the TV and newspaper readers in showing a positive attitude. Age difference and education level may be considered as an essential factor for choosing different media as a source for information during this time (Olum et al., 2020).

## 5. Limitations

COVID-19 is a novel virus that has been identified just before the onset of 2020. We faced difficulties in interpreting the results as there is no single KAP study in the food industries. As it was an online-based study and self-reported data were used, there is a possibility of biasness from the respondents. People from the supply chain department could be included in this study. The supply chain department plays a significant role in raw materials safety. Due to some limitations, we failed to arrange people from there. It would be more fruitful if we could observe the changes in their KAP over time.

## 6. Conclusion and recommendations

The article approaches a topic that may be of great interest to workers, consumers, processing plant managers, as well as public health systems and their evolution, in order to ensure improvement, particularly when it comes to the interaction between these systems and public policies in the considered area. According to the findings, food handlers of Bangladeshi food industries have adequate knowledge, a positive attitude, and desired practices regarding the COVID-19 issue. Education and work experience showed to have a positive impact on the KAP. It is critical to consider the importance of hiring employees with a higher level of education as well as providing necessary and regular training. For the current pandemic situation, a KAP study could be useful in determining the effectiveness of any COVID-19-related training for industry personnel. In the perspective of Bangladesh, more extensive research on the COVID-19 KAP should be conducted among food handlers in restaurants, street-food shops, and small and medium-sized businesses (SME), because people in this country usually buy foods from these sources more frequently than from food industries. Furthermore, additional research may be conducted to determine the KAP differences and significance of training among trained and untrained employees in various food and pharmaceutical industries. The current study will hopefully aid researchers and policymakers in developing their study and questionnaire for conducting respective research.

## Data Availability

All data produced in the present study are available upon reasonable request to the corresponding author.

## Acknowledgement

We especially thank the section managers for their cordial help. We also would like to appreciate the food handlers for their voluntary participation in this study.

## Data availability statement

Data will be available upon reasonable request.

## Funding information

There has been no funding and financial support for this study.

## Declaration of competing interest

We declare that we have no conflict of interest.

## Notes

### Competing Interest Statement

The authors have declared no competing interest.

### Funding Statement

No funding received

### Author Declarations

The institutional review committee of Sylhet Agricultural University, Bangladesh reviewed the study materials and approved the study.

### Summary of Updates

1. Simple modifications in the title, abstract, and introduction. 2. Ethical statement has been added within the manuscript. 3. The current version consists more data and further analysis.

